# Sex and Gender Influences on Problematic Cannabis Use and Cannabis Use Disorder: A Scoping Review Protocol

**DOI:** 10.1101/2025.06.21.25330052

**Authors:** Justin Matheson, Catharine A. Mielnik, Rodney Knight, Madison Wright, Noa Marley, Sarah Bonato, Bernard Le Foll, Louise Gallagher, Dafna Sara Rubin-Kahana

## Abstract

**Introduction:** Cannabis use and cannabis use disorder (CUD) are more prevalent among men and boys than among women and girls. However, this sex/gender gap has been narrowing in recent decades, likely due to an increase in cannabis use among women and girls, who have been historically under-represented in cannabis research. The lack of sex- and gender-based approaches within cannabis research has been highlighted in previous reviews, some of which have synthesized existing literature of associations between sex, gender, and cannabis use. What is missing is a clinically-relevant synthesis of evidence for sex and gender influences on problematic cannabis use, including treatment-related outcomes that could be used to influence care. The objective of this scoping review is to identify and synthesize published evidence about the influence of sex and gender on correlates and outcomes of treatment among people with problematic cannabis use (including CUD). Furthermore, we will examine to what extent this published literature has considered how sex and gender intersect with other social categories such as race and sexuality.

**Methods and Analysis:** This scoping review will follow the most commonly used methodology, the 2005 Arksey and O’Malley scoping study framework (including the optional consultation exercise to solicit feedback from relevant stakeholders) and the Preferred Reporting Items for Systematic reviews and Meta-Analyses (PRISMA) Extension for Scoping Reviews reporting guidelines. We will search MEDLINE, Embase, CINAHL, PsycINFO, and Web of Science for articles published between 2010 and the present. Included studies must be conducted in human participants with problematic cannabis use (e.g., diagnosis or screening for CUD) and include an analysis of sex- and/or gender-related factors. Using Covidence software, two independent reviewers will screen each record at the title/abstract and full text phases. Two independent reviewers will then use a data charting form developed by the study team to extract data. Data charting and both phases of article screening will begin with a pilot process completed by the entire team to ensure consistency. Article data will be exported into a spreadsheet to facilitate summary and basic descriptive statistics. Studies will be grouped together first by content area (e.g., treatment correlates, treatment effectiveness), then by study design, and which sex- or gender-related factors are considered in the analysis.

**Dissemination:** We will disseminate findings using two main strategies. First, we will engage in traditional knowledge translation, including publication in peer-reviewed journals and presentation at both medical and scientific conferences. Second, we will engage in knowledge translation strategies that will reach a wider audience (e.g., presentations to non-researcher audiences, dissemination of findings through social media networks, and development of brochures, infographics, and short videos to summarize our findings for a lay audience). We aim to ultimately engage relevant stakeholders (including clinicians) to determine how the identified evidence can best support care of problematic cannabis use.

## INTRODUCTION

### Background and Rationale

According to the United Nations Office on Drugs and Crime (UNODC), 228 million adults worldwide used cannabis in 2022 (1). There is a historical sex/gender gap in the prevalence of cannabis use, where men and boys are consistently more likely to use cannabis than women and girls across different metrics of use (e.g., lifetime use, past-month use, daily use) and in many countries globally (2-4). We refer to this as a “sex/gender” gap, as differences in cannabis use are likely driven by both biological and sociocultural factors (2). Importantly, the sex/gender gap seems to be narrowing. For example, the gap in past-month prevalence of cannabis use in the United States (US) declined from a high of 2.125 men to women using cannabis in 2002 to a low of 1.25 men to women in 2020 – which seems to be largely driven by an increase in women’s cannabis use (5). Youth data from the Canadian province of Ontario show that, as of 2023, girls actually have a higher prevalence of cannabis use than boys do (6). Changes in the sex/gender gap in prevalence of use may be driven in part by cannabis legalization and will likely lead to changes in cannabis-related harms, such as prevalence of cannabis use disorder (CUD) (2). CUD is characterized by an inability to cease or reduce cannabis use despite persistent physical and/or psychological harm (7), as defined in the most recent edition of the Diagnostic and Statistical Manual of Mental Disorders (DSM-5). While men are more likely to meet criteria for CUD and have greater symptom severity (8-11), women may escalate their use of cannabis faster than men (e.g., fewer years between age of first use and CUD symptom) (8, 10, 12, 13). Recent data-driven work has suggested that there are sex/gender differences in the factors underlying CUD – individual risk factors (e.g., mental health or cognitive factors) had stronger associations with CUD in men, while social-structural risk factors (e.g., low education level) had stronger associations in women (14). More work is needed to better understand how sex and gender impact CUD trajectories, correlates, and treatment and how to integrate this knowledge clinically to improve care for individuals with CUD.

Sex is an umbrella term that refers to complex biological traits and processes necessary for sexual reproduction, including genes and gene expression, hormones, body parts, and all the downstream biological impacts of sex-related gene expression and hormone production. Sex is thus a property of bodies and affects the way that drugs interact with bodies (3). A growing literature has found that gonadal hormones like estrogens, progestins, and androgens (which are essential for sex-related biological processes) can influence drug-seeking behaviours in rodents and the experience of subjective drug states in humans (15-18). For example, female rodents show greater self-administration of cannabinoids than male rodents, yet this sex difference is reduced by ovariectomy (removal of the ovaries, and thus dramatic reduction in ovarian hormone production) (19). The modulation of drug experiences by ovarian hormones is thought to contribute to variability in the risk of progression of drug use in humans (20). Moreover, human laboratory experiments have found that adult female participants experience greater subjective effects of cannabis than male participants under certain conditions (21-23).

Gender is a term that describes the sociocultural context and correlates of sexed bodies – it is a social construct that describes the social meaning of sexed bodies and the organization of that meaning into socially relevant categories. Gender is typically used as an umbrella term to refer to multiple concepts, including gender identity (individual experiences of gender), gender roles and norms (expectations placed on individuals based on their presumed gender), gender relations (how individuals of different genders interact, especially as it relates to power), and institutional gender (the translation of gendered meaning into social institutions, e.g., laws and regulations dictating the distribution of power and resources) (3). Gender roles and norms can influence drug use and harms, e.g., stigmatization and devaluation of women’s drug use partially explains lower drug use and more experiences of sociocultural harms associated with persistent drug use among women (2). The historical association between substance use and masculinity (24) has likely had an important role in shaping sex/gender differences in cannabis use, where men and boys have a higher prevalence of use than women and girls across cultures (2). Performative aspects of gender (i.e., engaging in behaviours to shape or reinforce one’s gender identity or expression) can influence how and why drugs are used and in which contexts (3). In quantitative studies, greater adherence to traditional masculine norms is associated with increased use of cannabis, while adherence to feminine norms is associated with decreased use of cannabis (2, 25). Several studies have focused on initial opportunities to use cannabis, finding that boys can more easily access cannabis than girls (2).

Some nuances about sex and gender are worth noting. First, the scientific literature is increasingly acknowledging that neither sex nor gender are binary (26, 27). Further, there is a distinction between an individual’s gender identity (e.g., woman, man, or non-binary) and their expression or performance of gender (e.g., femininity and/or masculinity) (26, 27). Second, there is an increasing focus on using an intersectional framework in the health sciences to understand how multiple social categories (e.g., gender, race, sexuality) intersect at the individual level to influence health (28). For example, while sexual and gender minorities have a higher prevalence of cannabis use than cisgender, heterosexual people, bisexual women appear to have the highest prevalence of use when examining intersections of gender and sexual identity (29). Complex three-way intersections between gender, sexual, and racial identities have been documented for lifetime substance use (30). To the extent possible, our review will keep these nuances in mind when synthesizing and interpreting the results of the identified literature.

There are no currently approved pharmacological treatments for CUD, so CUD treatment primarily involves psychological and psychosocial interventions (31). The current evidence favours both motivational enhancement therapy (MET) and cognitive behavioural therapy (CBT) for treatment of CUD, while contingency management has shown promise in some studies as well (32). Gender likely influences psychosocial interventions. For example, previous studies have found that women with anxiety disorders attend more psychotherapy sessions, are more engaged in therapy, and view psychotherapy as more helpful when compared to men (33). Meta-analysis has identified that women with psychiatric disorders are more likely to prefer psychotherapy over pharmacotherapy when compared to men (34). Pharmacological interventions for CUD have been tested in clinical trials, with the strongest evidence in favour of drugs that target the endocannabinoid system, though the evidence is still weak and more randomized controlled trials are needed to establish efficacy (31). Importantly, there are sex differences in the endocannabinoid system. For example, circulating levels of endocannabinoid ligands fluctuate across the rodent estrous cycle (a hormonal cycle analogous to the human menstrual cycle) (35) and ovariectomy causes a change in density of the cannabinoid type-1 receptor (CB1R) that is reversed by administration of exogenous estradiol (36). Thus, both sex and gender have the potential to influence various CUD treatment approaches. Furthermore, sex/gender differences in comorbidities have the potential to influence CUD treatment. For example, studies have consistently documented a stronger association between CUD and poorer mental health among women than among men, particularly mood and anxiety symptoms (37-41).

Previous narrative reviews have highlighted broad relationships between sex, gender, and cannabis (e.g., 2, 15). Systematic reviews have each provided a specific piece of the puzzle – how sex interacts with cannabis use to impact the brain (42), changes in the sex/gender gap in the prevalence of cannabis use (43), and sex differences in the comorbidity of CUD with mental illness (44). Finally, a large scoping review was undertaken pre-2020 that broadly covered the influence of sex and gender on cannabis use and related outcomes (3, 25), including one paper that specifically focused on sex- and gender-based analysis in trials of pharmacological interventions for CUD (45). However, what is currently missing is a structured overview of all clinically relevant literature relating sex, gender, and intersecting social categories to problematic cannabis use and CUD to inform prevention and treatment of CUD. We chose a scoping review to account for the breadth of evidence that is necessary to be clinically meaningful, and to help us organize the existing evidence to identify significant knowledge gaps necessary to advance care. We chose to use the Arksey and O’Malley (2005) framework for scoping reviews (46) as this is the most commonly used scoping review methodology (47).

### Objective

The overarching objective of the present scoping review is to provide a synthesis of all published evidence (from 2010 to present) implicating sex and gender in problematic cannabis use and CUD treatment, trajectories, and correlates. Furthermore, across research questions, we will consider the scope of evidence for intersections of sex and/or gender with other social categories such as race, ethnicity, sexuality, and ability.

<u>Primary research question</u>: What is known from existing published evidence about the influence of sex, gender, and intersecting social categories on correlates and outcomes of treatment among people with problematic cannabis use?

We further break this research question down into three secondary questions:

1. How do sex and gender influence antecedents to treatment or correlates of treatment seeking (e.g., cannabis use patterns, symptom presentation or severity prior to entering treatment; motives for treatment seeking; barriers or facilitators to treatment seeking) among people with problematic use?
2. Are there sex/gender differences in mental health or substance use health characteristics of people currently in treatment for problematic cannabis use?
3. Are there sex/gender differences in treatment effectiveness or retention?

## METHODS

This scoping review protocol has been registered on Open Science Framework (OSF) Registries (Registration DOI: https://doi.org/10.17605/OSF.IO/KC76M). We will follow the scoping study methodology created by Arksey and O’Malley (46). This methodology involves five main steps: creating a research question, identifying relevant studies, selecting studies, charting data, and collating, summarizing, and reporting the results (46). An optional sixth component is the consultation exercise (consulting with relevant stakeholders, such as clinicians or clinical managers), which we will likely implement later on in the scoping review process (i.e., once all articles are screened and data is extracted). The scoping review protocol was written following guidelines provided by the Center for Open Science (48). We will follow the Preferred Reporting Items for Systematic reviews and Meta-Analyses (PRISMA) Extension for Scoping Reviews reporting guidelines (49).

### Eligibility Criteria

Inclusion criteria:

1. The article includes data from any study involving human participants (including observational and interventional designs) from any geographical location or sociocultural context. There will be no restrictions on age, race/ethnicity, or comorbidities. Studies will be excluded if they contain data from only non-human animal models or tissues/samples from humans with no whole-person data.
2. The study includes a group of human participants who meet at least one of two criteria: (i) Meets diagnostic or screening criteria for problematic cannabis use (i.e., DSM-5 Cannabis Use Disorder, DSM-4 Cannabis Abuse or Dependence, ICD-10 Cannabis Abuse or Dependence, or equivalent) based on self-report, clinician assessment, or screening above threshold in a validated scale (e.g., the Cannabis Use Disorders Identification Test [CUDIT] (50)); OR (ii) Are seeking treatment, in treatment, or have previously been treated for problematic cannabis use (e.g., reduction or cessation of cannabis use).
3. The study sample must meet one of two criteria related to sex/gender: (i) The study includes participants of at least two sex or gender categories (e.g., male and female youth; cisgender and transgender adults) and includes disaggregation or comparison of participant data by sex (e.g., sex at birth) or gender (e.g., gender identity); OR (ii) The study includes one sex or gender group, with at least one specific sex- or gender-related measure (e.g., a study measuring hormone levels in a single-sex group or assessing pregnancy-related outcomes in cisgender women). Studies will be excluded if they do not disaggregate data by sex or gender or if they contain just one sex or gender category without any consideration of sex- or gender-related factors.
4. Any outcome measures related to treatment for problematic cannabis use (e.g., DSM-5 CUD). This includes clinically relevant characteristics of people meeting diagnostic or screening criteria for problematic use. Any studies not including outcomes that have any direct relevance to treatment will be excluded.
5. The article is a primary, peer-reviewed research article available in English and was published after the year 2010. The year 2010 was selected because sex and gender concepts evolve rapidly, so we wanted to include mostly recent findings. Moreover, a 15-year period for article searches is in line with suggestions from prior methodological reviews (46, 49). Secondary sources (including reviews), articles not available in English, and articles published before 2010 will be excluded. We will include relevant grey literature that is completed but unpublished primary research, e.g., conference presentations, dissertations, and preprints.

In the event of any ambiguous situations that arise during title/abstract screening (e.g., an article missing an abstract that is not clearly ineligible based on the title), articles will automatically be moved on to full-text screening. This is particularly important for our review, as sex and gender terminology is often not included in the title or abstract of articles that would meet our eligibility criteria. This means that we will not apply criterion 3 at the title/abstract screening stage. At the full-text screening phase, any ambiguous situations will be brought to the full authorship team to discuss, and decisions will be documented in a table, which can be published as supplementary material with the final review article.

### Information Sources and Search Strategy

The draft of the Medline search strategy was developed by a research librarian at a large tertiary care mental health and addictions teaching hospital. The strategy was developed in collaboration with the review team and facilitated through email communication and regular virtual meetings. Starting with text mining previously identified on topic articles, a multiphase search approach was utilized to validate the search; numerous preliminary searches were performed to determine the suitability of the search strategy and the article eligibility criteria, in order to optimize how to identify research focusing on sex and/or gender. The final step of validating the search was to ensure that all on topic articles identified by the text mining and preliminary searches were captured by the expanded final Medline search. For the final searches, the following databases will be searched: MEDLINE, Embase, CINAHL, PsycINFO and Web of Science for articles published between 2010 and to the present. There will not be any language or study type limits applied to the search results, in order to not filter out relevant research unintentionally.

Additionally, a focused grey literature search will also be completed, restricted to key sources of completed but unpublished primary research. Specifically, the grey literature search will be limited to dissertations, preprints and conference papers/proceedings. Resources for searching the grey literature will include ProceedingsFirst, PapersFirst, ProQuest Dissertations and Theses Globe and medRxiv. We will also manually search reference lists from related review papers and from articles included in our review. We will perform the search just once. If sufficient time has elapsed, we may revisit this decision and update the registration as needed. We will use EndNote 21 (Clarivate, Philadelphia, PA, United States) for reference management. The Peer Review of Electronic Search Strategies (PRESS) checklist (51) will be used to document peer review of the search strategy.

The full search strategy for MEDLINE is provided here as an example:

**Database: Ovid MEDLINE(R) ALL <1946 to November 19, 2024>**

**Search Strategy:**

1. exp Marijuana Abuse/
2. ((cannabis* or marijuana* or marihuana* or hash? or hashish* or ganja or ganjas or bhang or bhangs or hemp or hemps) adj3 (abus* or addict* or dependen* or disorder* or harm? or harmful* or misus* or non-medical or nonmedical or recreat* or problem*)).mp.
3. exp Sex Factors/
4. exp sex characteristics/
5. exp sex distribution/
6. (sex* adj2 (factor* or characterist* or differ* or distribut*)).mp.
7. ((female* or male*) adj2 (differ* or similar* or dissimilar* or contrast* or varianc* or variation* or variabilit* or divergen* or deviat* or polarit* or gap? or split? or disparit* or imbalanc* or uneven* or even* or incongruit* or contradic* or contradistincti* or nonconform* or conform* or unlikeness or likeness or contrarie* or dissimilitud*)).mp.
8. ((women? or woman? or men? or man?) adj2 (differ* or similar* or dissimilar* or contrast* or varianc* or variation* or variabilit* or divergen* or deviat* or polarit* or gap? or split? or disparit* or imbalanc* or uneven* or even* or incongruit* or contradic* or contradistincti* or nonconform* or conform* or unlikeness or likeness or contrarie* or dissimilitud*)).mp.
9. exp Women/
10. exp Men/
11. wom#n?.mp.
12. m#n?.mp.
13. female*.mp.
14. male*.mp.
15. (9 or 11 or 13) and (10 or 12 or 14) [women/men/female/male]
16. (male* or men? or man? or boy or boys).mp.
17. (female* or women? or woman? or girl or girls).mp.
18. 16 not 17 [citations mention men/males but not women/females]
19. 17 not 16 [citations mention women/females but not men/males]
20. gender*.mp.
21. exp animals/ not humans.sh.
22. 1 or 2 [cannabis misuse set]
23. (or/3-8) or 15 or 18 or 19 or 20 [gender set]
24. exp animals/ not humans.sh.
25. (22 and 23) not 24
26. limit 25 to yr=“2010-Current”

### Study Selection and Screening

Duplicate records will be removed using Endnote or Covidence software before beginning the screening process. Eligibility criteria will be applied at two stages to determine article inclusion: (i) title/abstract screening; and (ii) full-text screening. Note that, as described earlier, not all eligibility criteria will be applied at the title/abstract screening stage in order to include as many relevant articles as possible (e.g., articles that do not mention sex or gender in the title or abstract but are otherwise eligible will move to full-text screening). Covidence systematic review software (Veritas Health Innovation, Melbourne, Australia; available at www.covidence.org) will be used for article screening and data charting. Two independent reviewers will screen each record at both stages. In the event of voting discrepancies between reviewers, we will first try to reach consensus between the original two reviewers; if this is not possible, we will discuss any discrepancies with the review team, including at least one of the first author and senior author, to reach a final decision.

Both stages of article screening will begin with a pilot process (sometimes called a calibration exercise (49)). The research librarian will generate a random sample of records (50 for the title/abstract stage, 10 for the full-text stage) and the entire team will screen these records in Covidence. Once the pilot process is completed, the team will discuss the review process and any discrepancies between reviewers. If there is lower than 80% proportionate agreement between any two reviewers (calculated in Covidence), the screening process will be reviewed, any necessary adjustments will be made, and the team will perform a second pilot process. Screening at both stages will proceed only once the pilot process is completed and all team members are in agreement with the screening process and feel they have adequate training to begin. We will use the pilot process as an opportunity to determine an appropriate pace for screening (i.e., we will decide on a weekly goal for number of articles reviewed per reviewer for each stage of screening).

Given the broad nature of the evidence we plan to synthesize in this scoping review, it is likely that we may be required to make changes to the study selection process (e.g., updating or revising eligibility criteria). Any changes made will be documented in a log and will be published along with the final review article (likely as supplementary material).

### Data Charting

The data charting form will be created with input from the entire authorship team. Two independent reviewers will extract data using Covidence software. If there are any discrepancies in data extraction, we will follow the same process as described for article screening. Missing, incomplete, or unclear data will be coded accordingly (e.g., we will code “Not Reported” if the data are not included in the article or “Unclear” if the team cannot reach a consensus on how to interpret the data including in the article). The team will make attempts to contact the corresponding author of articles containing any missing or unclear data. If the corresponding author replies with the requested information, this will be updated in the charting form for that article. We will keep a log of author contacts to be transparent about data that was not included in the published article. If the corresponding author has not replied to the study team after one initial email and two follow-up emails, or if the contact information for the corresponding author is no longer valid, we will leave the data coded as “Not Reported” or “Unclear”. If we encounter any “friend studies” (i.e., multiple articles published from the same study), we will include them all if they contain separate relevant endpoints, or else we will use the article with the largest sample size and/or longest follow-up period for data extraction. We will not be conducting a critical appraisal or risk of bias analysis, given the anticipated heterogeneity in study design.

We will begin the data charting with a pilot process. The research librarian will randomly select 10 articles, and the entire team will participate in pilot data charting. Once this is complete, the team will meet to discuss the data charting process and any discrepancies. Data charting will proceed only once the pilot process is completed and all team members are in agreement with the data charting form and feel they have adequate training to begin. Regular team meetings will be held with at least one of the first or senior author present to discuss any issues that arise during data charting. Any changes made to the data charting form will be documented in a log and will be published along with the final review article (likely as supplementary material).

The following data elements will be extracted from each article:

- Title of paper
- Author(s)
- Journal title
- Year of publication
- Year(s) of data collection
- Study location
- Study design
- Definition of sex or gender (e.g., sex at birth, gender identity, or “absent” if no definition is provided)
- Intersecting social identities, if applying an intersectional framework
- Sex/gender conceptualization or theoretical framework (e.g., the authors assume a biological association between sex at birth and relevant outcomes, or include a specific theoretical model of gender)
- Sample size (including breakdown by sex or gender, where relevant)
- Age range and/or mean and standard deviation
- Cannabis use characteristics of the sample (e.g., diagnostic criteria applied, CUDIT scores)
- Primary study objective
- Relevant outcome measures
- Method for measuring each relevant outcome
- Results
- Author conclusions

### Synthesis and Presentation of Results

All extracted data will be reviewed by a third member of the review team not originally involved in data extraction. In order to summarize data and present basic descriptive statistics, data will be exported into a spreadsheet in Microsoft Excel (Microsoft Corporation, Redmond, WA, United States). A PRISMA flow diagram will be used to graphically depict the flow of article capture, screening, and extraction (52).

Articles will first be grouped together with regards to the specific research question they best inform (1, 2, or 3, as described earlier). Within each group, subgroups will be created based on study design and whether the analysis or theoretical framework considers biological (i.e., sex-related) or sociocultural (i.e., gender-related) factors. Depending on the results we identify, we may further group by age (e.g., results for youth separate from adults). We will likely conduct a consultation exercise at this phase, in order to solicit feedback and perspectives from relevant stakeholders (e.g., clinicians) related to the study results. We will use tables to present relevant data for each article included in each of these subgroups. Descriptive analysis will be used to complement these tables and communicate the frequencies of each type of study.

### Ethics and Dissemination

This scoping review is exempt from ethics review as we will not be directly accessing any data.

Multiple members of the review team are clinician-researchers, who are well situated to both inform the review process and guide dissemination of the results to have maximum capacity to influence policy and care. The review team has extensive experience engaging with knowledge translation (KT) services within our primary institution (CAMH). Our standard practice is to engage KT efforts on two fronts. First, we engage in traditional KT, such as publication in peer-reviewed journals and presentation at both medical and scientific conferences. Second, we will work with internal services (such as media affairs and patient and family engagement services) to develop broader KT efforts that will reach a wider (non-academic) audience. This can include KT activities such as presentations to broad audiences through our Patient and Family Learning Space, dissemination of findings through social media networks, and development of brochures, infographics, and short videos to summarize our findings for a lay audience. Our specific KT plan will depend on the scope of the evidence we generate in this review. We may consider bringing on additional collaborators (e.g., clinicians or other knowledge users) to assist in dissemination of findings to additional networks not covered by the current team.

## CONCLUSION

The goal of this scoping review is to identify and synthesize published evidence about the influence of sex and gender on correlates and outcomes of cannabis-related treatment among people with problematic cannabis use (including CUD). Furthermore, we want to characterize sex/gender differences in antecedents to treatment or correlates of treatment-seeking, characteristics of people in CUD treatment, and CUD treatment effectiveness. We will consider intersections of sex and/or gender with other social categories (e.g., race, sexuality) throughout the review. We hope this will serve as a useful evidence map to inform and advance treatment and prevention of CUD through a sex/gender lens. We also hope to stimulate future systematic reviews and meta-analyses that can more directly inform evidence-based care of CUD.

## Data Availability

No datasets were generated or analysed during the current study. All relevant data from this study will be made available upon study completion.

